# Causal Associations of Self-Reported Walking Pace with Telomere Length in 405,981 middle-aged adults: a UK Biobank study

**DOI:** 10.1101/2021.09.06.21263163

**Authors:** Paddy C. Dempsey, Crispin Musicha, Alex V. Rowlands, Melanie Davies, Kamlesh Khunti, Cameron Razieh, Iain Timmins, Francesco Zaccardi, Veryan Codd, Christopher P. Nelson, Tom Yates, Nilesh J Samani

## Abstract

**Objectives:** Walking pace is a strong marker of functional and health status. We investigated whether walking pace is also associated with leucocyte telomere length (LTL), which is causally associated with several diseases and has been proposed as a marker of biological age.

**Methods:** We used baseline data from UK Biobank participants recruited from March-2006 to July-2010. Walking pace was self-reported as slow, steady/average, or brisk. Accelerometer-assessed measures of total physical activity and intensity were included to support interpretation of walking pace data. LTL was measured by qPCR assay. Bi-directional Mendelian randomization (MR) analyses were undertaken to inform likely causal directions.

**Results:** The analysed cohort comprised 405,981 adults (54% women) with mean age of 56.5 years (SD, 8.1) and body mass index 27.2 kg/m^2^ (SD, 4.7). Steady/average and brisk walkers had significantly longer LTL compared with slow walkers, with a Z-standardised LTL difference of 0.066 (0.053-0.078) and 0.101 (0.088-0.113), respectively. Associations remained but were attenuated following full covariate adjustment: 0.038 (0.025-0.051) and 0.058 (0.045-0.072), respectively. Accelerometer data (n=86,002) demonstrated a non-linear association between LTL and habitual movement intensity, but not total activity. MR analysis supported a causal association of walking pace on LTL, with an increase in Z-standardised LTL of 0.192 (0.077, 0.306) for each difference in walking pace category. No evidence of a causal association was observed for LTL on walking pace.

**Conclusion:** Faster walking pace may be causally associated with longer LTL, which could explain some of the beneficial effects of brisk walking on health status.

## INTRODUCTION

Walking is a simple and accessible form of physical activity (PA) for all ages, conferring many physical, mental, and social health benefits with minimal adverse effects [1-4]. It therefore holds strong potential as a pragmatic target for intervention [5]. Strong associations with health status have been seen for habitual or self-rated walking pace, which has been associated with better physical fitness and reduced risk of cardiovascular disease and all-cause mortality [6-10], with brisk walkers having up to 20 years greater life expectancy compared to slow walkers [11]. Indeed, walking pace has been shown to have a stronger association with survival and be a substantially better prognostic marker for all-cause or cardiovascular mortality than other measures of PA volume, diet, or physical function [12, 13]. Similarly, accelerometer-assessed measures of PA in UK Biobank suggest that as little as 10 min of brisk walking a day is associated with longer life expectancy [14]. A genome-wide association study (GWAS) on self-reported walking pace within UK Biobank identified 70 independent SNPs at genome-wide significance [15], close to an order of magnitude greater than the number reported for other self-reported or accelerometer-assessed measures of PA traits within the same cohort [16].

The importance of walking pace as a marker and potential promoter of health is likely to reflect it being a complex functional activity influenced by many factors, such as motor control, musculoskeletal health, cardiorespiratory fitness and lung capacity, habitual activity levels, cognition, motivation, and mental health [3, 13, 15]. These factors also align to the concept of biological age, which relates to an individual’s ability to maintain a robust homeostasis when subject to stressors [17]. Therefore, it is possible that walking pace acts as both a marker and modulator of biological age. However, whether walking pace is causally associated with potential indicators of biological age remains unknown.

Although the relationship between leukocyte telomere length (LTL) and disease is complex [18], LTL has been proposed as a marker of biological age and is associated with higher risk of several age-related diseases; including coronary artery disease and several cancers [18-23]. Telomeres are DNA–protein complexes that protect the ends of chromosomes from degradation, end-to-end fusion, and abnormal recombination of DNA strands (genomic instability). The DNA component progressively shortens with each cell cycle, decreasing in most cell types as humans age, ultimately contributing to replicative senescence [19, 24]. Along with reflecting cellular replicative history, telomere shortening is also moderated by factors such as oxidative stress and inflammation [24]. Telomere length is usually measured in leukocytes (LTL), which is reflective of telomere length in other tissues, along with reflecting the senescent status of circulating cells related to the immune system [25].

Previous research suggests an association of higher levels of PA and cardiorespiratory fitness with longer LTL [26, 27], supporting the hypothesis that higher levels of PA and cardiorespiratory fitness may act to slow markers of biological ageing. However, most studies in humans to date have been small and/or observational in nature, with some studies showing weak or null associations [26-28]. Therefore, the current literature is not definitive and does not support inferences around causal direction. Moreover, there remains insufficient research investigating the association between simple functional habitual movements, such as walking pace, and LTL. The aim of this study was, therefore, to investigate the association between self-reported walking pace and LTL in middle aged adults. This included harnessing previously defined genetic instruments for both walking pace and LTL to undertake bi-directional Mendelian randomisation (MR) analyses, to help clarify the causal nature and relative importance of any observed associations. We support observational analyses for self-reported walking pace using accelerometer-assessed total PA and intensity, to aid broader interpretation. Our observational hypothesis was that a brisker walking pace would be causally associated with longer LTL.

## METHODS

### Data source and study population

This analysis used data from participants within UK Biobank, a large prospective cohort of middle-aged adults designed to support biomedical analysis focused on improving the prevention, diagnosis, and treatment of chronic disease through phenotyping and genomics data [29]. Between March 2006 and July 2010, individuals living within 25 miles of one of the 22 study assessment centres located throughout England, Scotland, and Wales were recruited and provided comprehensive data on a broad range of demographic, clinical, lifestyle, and social outcomes. All participants provided written informed consent and the study was approved by the NHS National Research Ethics Service (Ref: 11/NW/0382).

### Self-reported walking pace

Self-reported walking pace was ascertained using a touchscreen question: “How would you describe your usual walking pace?” with response options of “slow”, “steady/average” or “brisk”. Participants could access further information which defined a slow pace as less than 3 miles per hour, a steady/average pace as between 3-4 miles per hour, and a brisk pace as more than 4 miles per hour.” We excluded participants whose answers were “None of the above or “Prefer not to answer” (n = 3,956).

### Covariate measurement

We utilized demographic and lifestyle related characteristics of age, sex, ethnicity (white/non-white), Townsend Index of deprivation, highest educational level achieved (degree or above/any other qualification/no qualification), employment status (unemployed/in paid or self-employment), alcohol drinking status (never/previous/current), salt added to food (never/sometimes), oily fish intake (never/sometimes), fruit and vegetable intake (a score from 0-4 taking into account questions on cooked and raw vegetables, fresh and dried fruit consumption), processed and red meat intake (average weekly frequency in days per week), and sleep duration (<7, 7-8, >8 hours), and a diagnosis of cardiovascular disease or cancer prior to baseline. The latter two prevalent disease variables were derived from the self-reported history of heart attack, angina, stroke, or cancer variables, and from linked hospital episode data (corresponding ICD 10 codes I20-25, I60-69, or C00-99). Health-related covariates of blood pressure and cholesterol medication, doctor diagnosed diabetes or prescribed insulin medication and mobility limitations (self-reported longstanding illness or disability or chest pain at rest), white blood cell (leukocyte) count, and body mass index (BMI) in three categories (<25, 25-30, ≥30 kg/m^2^) were included in models. Total MET-minutes/week of PA was derived from weekly frequency and duration of walking, moderate, or vigorous intensity PA using the short-form International Physical Activity Questionnaire (IPAQ) [30]. Further details for each variable are available on the UK Biobank Website https://www.ukbiobank.ac.uk/

### Accelerometer PA measurements

PA was assessed using accelerometry in a subset of participants (n=∼100,000; see **Supplemental Figure S1**) between 2013-2015 who were invited to wear an Axivity AX3 triaxial accelerometer (Axivity Ltd., Newcastle, UK) continuously on their dominant wrist for seven consecutive days [31]. Accelerometer measures were average acceleration over the 24 h day (proxy for total PA, m*g*) and intensity gradient over 24 h (a measure of the intensity distribution of PA over the day) [31-33]. A higher average acceleration indicates more PA is accumulated across the day, irrespective of the intensity. A higher intensity gradient indicates more time is habitually spent in higher intensity activities, such as brisk walking (see **Supplemental Accelerometer Methods** for the accelerometer data processing methodology and PA variable interpretation).

### Leucocyte telomere length measurements

LTL was measured using an established multiplex qPCR assay from 488,415 available DNA samples of participants in UK Biobank, which are detailed elsewhere [34]. After extensive quality checks and adjustment for technical factors valid LTL measurements were available for 472,577 individuals [34]. For analyses of data available for the full cohort we used log-transformed and z-standardised LTL values (UK Biobank data field code 22192), of which log-transformed data were re-standardised for analyses performed on the sub-set of participants with accelerometer data.

### Statistical analyses

For analyses presented in this paper, we included all participants with LTL measured from the UK Biobank baseline sample, where there was no mismatch in self-reported and genetic sex (n=472,248). Exclusions were also made for missing walking pace or covariate data, or for missing accelerometer data in the subset analysis (**Supplemental Figure S1**).

A series of linear regression models were used to quantify the associations of “steady/average” and “brisk” self-reported walking pace with Z-standardised log-LTL (*β*-coefficient with 95% CI), compared to “slow” walkers as the reference group. Model 1 adjusted for age, sex, ethnicity and total white blood cell count, as these variables are known to be associated with LTL [34]. Model 2 additionally adjusted for other included confounders. Models 3 and 4 additionally adjusted for total PA and then body mass index, which were considered last given their potential role as confounders or mediators.

#### One-sample bidirectional MR

We conducted a bi-directional one-sample Mendelian randomisation (MR) analysis (see **Supplemental Figure S2**) to evaluate a potential causal relationship between longer LTL and self-rated walking pace, using the inverse-variance weighted (IVW) [35] method with sensitivity analyses using both the weighted median [36] and robust adjusted profile score [37] methods. We also used MR-Egger regression to assess robustness to horizontal pleiotropy [38]. Each of these approaches makes a slightly different set of assumptions about the pleiotropic effects of genetic instruments, hence if the effect estimates are consistent across methods this provides stronger evidence of causality. To examine the causal association of LTL on walking pace (*part 1*) the LTL instrument utilised a set of 130 genome-wide significant (P<8.31×10^−9^), conditionally independent, uncorrelated, and non-pleiotropic genetic variants we recently identified as genetic instruments for LTL [18]. We matched variants to the publicly available walking pace GWAS data that were unadjusted and adjusted for BMI, matching 121 variants. For walking pace on LTL (*part 2*) we considered 70 genome-wide significant (P<5.0×10^−8^) independent genetic variants as the instrument from the unadjusted GWAS using weights from both the unadjusted GWAS and the BMI-adjusted GWAS [15]. These were matched to the LTL GWAS [18], matching all variants. To interpret the causal effect estimate of 1-SD increased LTL length on differences in walking pace category, the coded values 0, 1 and 2 for self-reported slow, steady/average and brisk walking pace can be thought of as threshold values for an underlying continuous trait, as has been demonstrated previously [15].

#### Sensitivity analyses

To support the findings and interpretation for self-reported walking pace, we included sensitivity analyses examining the sub-set of the UK Biobank cohort with accelerometer data and LTL (n=86,002; see **Supplemental Figure S1** and **Supplemental Table S1** for sub-sample descriptive characteristics), focussing on two key metrics summarizing total PA (average acceleration) and the intensity distribution (intensity gradient) of PA over each 24-hour day (see **Supplemental Accelerometer Methods**). Due to some evidence of non-linearity, associations for these two accelerometer exposures with LTL were examined using restricted cubic splines (three evenly-spaced knots), with reference values set at the 10^th^ percentile of the exposure.

Observational analyses were conducted using Stata v15.1 (StataCorp, TX, USA) and statistical significance was set at *p*<0.05 (two-tailed). MR analyses were performed using the *MendelianRandomisation* package implemented in R software [39].

## RESULTS

### Descriptive characteristics of the observational analytical sample

Descriptive characteristics for the analysis sample are shown in **Table 1**. Mean age was 56.5 years (SD, 8.1); mean BMI was 27.2 kg/m^2^ (SD, 4.65); and 54% and 95% were female and white, respectively. Approximately half the participants reported an average/steady walking pace (n=212,303; 52.3%), with 6.6% (n=26,835) reporting a slow walking pace and 41.1% (n=166,843) reporting a brisk pace. Compared to slow walkers, those who reported being average/steady and brisk walkers were slightly younger, were more likely to have never smoked, and were less likely to be taking cholesterol/blood pressure medications, have a chronic disease, or have mobility limitations. Slow walkers reported engaging in less PA and had a higher deprivation index and prevalence of obesity compared to average and brisk walkers. Differences observed between the walking pace groups in the overall sample were mostly comparable for the accelerometer sub-sample (**Supplemental Table S1**).

**Table 1.** Descriptive characteristics at baseline of the main analytical sample and by self-reported walking pace.

| Characteristics | Total sample | Slow | Average/steady | Brisk |
| --- | --- | --- | --- | --- |
|  | N=405,981 | N=26,835 | N=212,303 | N=166,843 |
| Age (years), mean (SD) | 56.46 (8.10) | 58.95 (7.51) | 57.14 (8.03) | 55.20 (8.09) |
| Female gender, n (%) | 217,267 (53.5%) | 14,444 (53.8%) | 113,832 (53.6%) | 88,991 (53.3%) |
| White ethnicity, n (%) | 386,820 (95.3%) | 24,279 (90.5%) | 200,811 (94.6%) | 161,730 (96.9%) |
| **Highest educational level achieved, n (%)** |  |  |  |  |
| *No qualification* | 63,306 (15.6%) | 8,603 (32.1%) | 38,022 (17.9%) | 16,681 (10.0%) |
| *Any other qualification* | 203,534 (50.1%) | 12,518 (46.6%) | 109,862 (51.7%) | 81,154 (48.6%) |
| *Degree level or above* | 139,141 (34.3%) | 5,714 (21.3%) | 64,419 (30.3%) | 69,008 (41.4%) |
| Townsend indicator of multiple deprivation, median (IQR) | -2.23 (-3.69-0.33) | -2.23 (-3.69-0.33) | -0.83 (-3.01-2.49) | -2.21 (-3.67-0.37) |
| In employment, n (%) | 237,656 (58.6%) | 237,960 (58.6%) | 8,414 (31.4%) | 119,230 (56.2%) |
| **Cigarette smoking, n (%)** |  |  |  |  |
| *Never* | 223,350 (55.0%) | 11,876 (44.3%) | 114,422 (53.9%) | 97,052 (58.2%) |
| *Previous* | 141,890 (34.9%) | 10,416 (38.8%) | 75,768 (35.7%) | 55,706 (33.4%) |
| *Current* | 40,741 (10.0%) | 4,543 (16.9%) | 22,113 (10.4%) | 14,085 (8.4%) |
| **Alcohol consumption, n (%)** |  |  |  |  |
| *Never or previous* | 29,449 (7.3%) | 4,480 (16.7%) | 15,824 (7.5%) | 9,145 (5.5%) |
| *< Twice a week* | 193,748 (47.7%) | 14,238 (53.1%) | 105,542 (49.7%) | 73,968 (44.3%) |
| *At least three times a week* | 182,784 (45.0%) | 8,117 (30.2%) | 90,937 (42.8%) | 83,730 (50.2%) |
| **Added salt intake, n (%)** |  |  |  |  |
| *Never/rarely* | 228,030 (56.2%) | 12,960 (48.3%) | 114,910 (54.1%) | 100,160 (60.0%) |
| *Sometimes or more frequent* | 177,951 (43.8%) | 13,875 (51.7%) | 97,393 (45.9%) | 66,683 (40.0%) |
| **Oily fish consumption, n (%)** |  |  |  |  |
| *More than once a week* | 229,701 (56.6%) | 13,923 (51.9%) | 116,979 (55.1%) | 98,799 (59.2%) |
| Fruit and vegetable intake score, median (IQR) | 2.00 (1.00-2.00) | 2.00 (1.00-2.00) | 1.00 (1.00-2.00) | 1.00 (1.00-2.00) |
| Weekly frequency of red or processed meat intake, median (IQR) | 0.75 (0.50-1.25) | 0.75 (0.50-1.25) | 0.88 (0.50-1.25) | 0.75 (0.50-1.25) |
| **Mean sleep duration, n (%)** |  |  |  |  |
| *<7 hours/day* | 97,302 (24.0%) | 8,492 (31.6%) | 50,447 (23.8%) | 38,363 (23.0%) |
| *7-8 hours/day* | 278,701 (68.6%) | 14,223 (53.0%) | 145,043 (68.3%) | 119,435 (71.6%) |
| *>8 hours/day* | 29,978 (7.4%) | 4,120 (15.4%) | 16,813 (7.9%) | 9,045 (5.4%) |
| **Body mass index, n (%)** |  |  |  |  |
| *Normal weight (<25 kg/m<sup>2</sup>)* | 137,660 (33.9%) | 4,214 (15.7%) | 57,872 (27.3%) | 75,574 (45.3%) |
| *Overweight (25-30 kg/m<sup>2</sup>)* | 174,368 (42.9%) | 8,854 (33.0%) | 95,206 (44.8%) | 70,308 (42.1%) |
| *Obese (≥30 kg/m<sup>2</sup>)* | 93,953 (23.1%) | 13,767 (51.3%) | 59,225 (27.9%) | 20,961 (12.6%) |
| Current prescription of blood pressure or cholesterol medicine, n (%) | 108,650 (26.8%) | 108,771 (26.8%) | 13,605 (50.7%) | 63,301 (29.8%) |
| Diagnosis of diabetes or insulin prescription, n (%) | 19,821 (4.9%) | 19,843 (4.9%) | 4,048 (15.1%) | 11,614 (5.5%) |
| Previous diagnosis of cardiovascular disease, n (%) | 25,887 (6.4%) | 26,214 (6.5%) | 5,400 (20.1%) | 14,566 (6.9%) |
| Previous diagnosis of cancer, n (%) | 33,250 (8.2%) | 33,965 (8.4%) | 3,207 (12.0%) | 18,077 (8.5%) |
| Mobility limitation, n (%) | 154,951 (38.2%) | 155,135 (38.2%) | 20,834 (77.6%) | 83,977 (39.6%) |
| Total MET-minutes/week physical activity, median (IQR) | 1668 (748-3372) | 1668 (748-3372) | 813 (330-1980) | 1583 (702-3272) |
| Total white blood cell (Leukocyte) count (10<sup>9</sup> cells/Litre), median (IQR) | 6.61 (5.61-7.80) | 6.61 (5.61-7.80) | 7.30 (6.15-8.67) | 6.73 (5.71-7.91) |
| Telomere length (z-score), mean (SD) | 0.004 (0.998) | 0.004 (0.998) | -0.133 (1.016) | -0.022 (0.998) |

Townsend score, a composite area-level measure of deprivation based on unemployment, non-car ownership, non-home ownership, and household overcrowding; a higher score indicates higher deprivation.

### Observational associations of walking pace and accelerometer-derived PA with LTL

The associations of walking pace with LTL are shown in **Figure 1**. For the minimally-adjusted model (model 1) steady/average and brisk walkers had significantly longer LTL compared to slow walkers: standardised difference 0.066 (95% CI: 0.053-0.078) and 0.101 (0.088-0.113), respectively. Associations for steady/average and brisk walking pace remained statistically significant, but were attenuated, following adjustment for potential confounding variables: [model 2; steady average = 0.038 (0.025-0.051) and brisk = 0.058 (0.045-0.072)]. Further sequential adjustment for total self-reported PA volume (MET-min/week) and BMI (models 3 and 4) did not materially alter the results (**Figure 1**).

**Figure 1.**
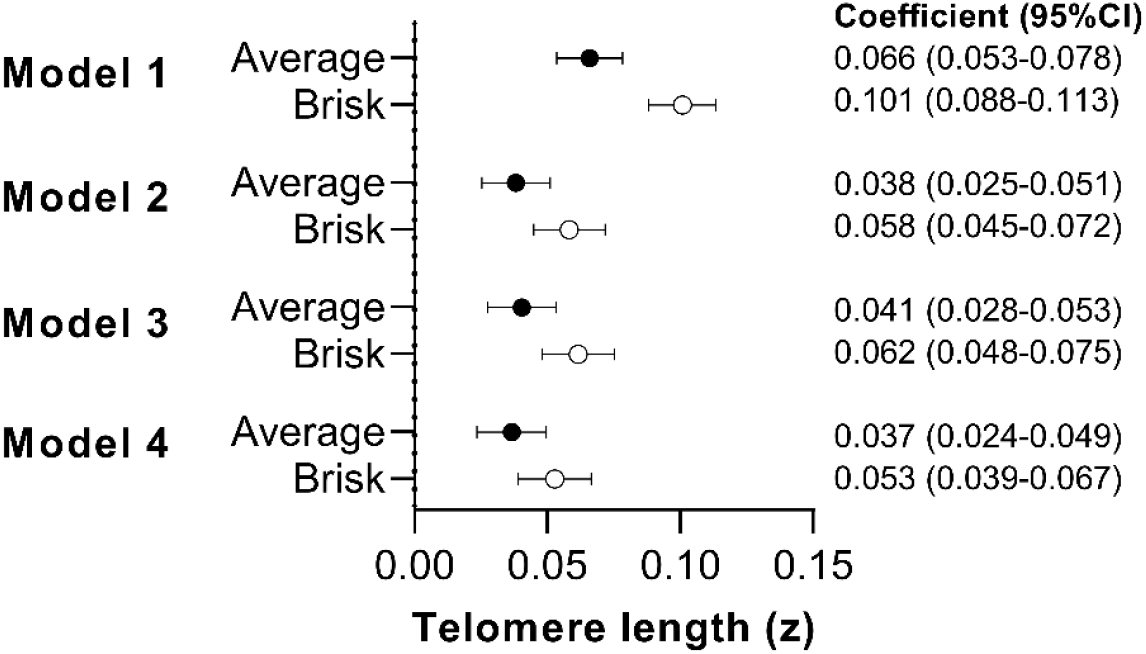
Associations between self-reported walking pace and LTL Data presented as *β*-coefficient (95% CI) for “average” (n=212,032) and “brisk” (n=166,641) walking pace relative to “slow” (26,804) walking pace (reference). Model 1: adjusted for age, sex, ethnicity, white blood cell count. Model 2: model 1 additionally adjusted for education level, employment status, Townsend index of deprivation, fruit and vegetable intake, processed and red meat intake, oily fish intake, regularity of adding salt to food, alcohol intake, smoking status, average sleep duration, blood pressure or cholesterol medication use, diabetes diagnosis or insulin prescription, mobility limitation, and prevalent cardiovascular disease and prevalent cancer. Model 3: model 2 additionally adjusted for total PA volume (MET-min/week). Model 4: model 3 additionally adjusted for body mass index.

### Accelerometer sensitivity analysis

Sensitivity analysis in the subset with accelerometer-derived continuous exposure measures of total PA and intensity (n=85,735) found that the intensity gradient had a positive non-linear association with LTL, showing that undertaking a greater proportion of daily PA at a higher intensity was associated with longer LTL, with associations retained (albeit attenuated) after covariate adjustment. In contrast there was little evidence of an association with total PA (**Supplemental Figure S3**).

### Bi-directional MR analyses

**Table 2** shows the MR analysis for the association of LTL on walking pace (part 1) and the association of walking pace on LTL (part 2), both with and without adjusting for BMI within the walking pace GWAS. There was no evidence of a causal association of LTL on walking pace, with or without adjustment for BMI. However, there was evidence that walking pace is causally associated with LTL. Per modelled difference in walking pace category (slow to steady/average, or steady/average to brisk) there was an increase in LTL SD of 0.192 (95% CI: 0.077, 0.306) before and 0.226 (0.061-0.388) after adjustment for BMI. No significant evidence of directional pleiotropy was found using MR-Eggers intercept (**Table 2**). Other MR methods showed broadly consistent findings; sensitivity analyses using weighted median MR attenuated the results somewhat, but the MR robust adjusted profile score estimated stronger effects.

**Table 2.** Mendelian randomisation between self-reported walking pace and LTL

| **Part 1: LTL on walking pace (121 SNPs for LTL)** |  |  |  |  |  |  |
| --- | --- | --- | --- | --- | --- | --- |
|  | **Unadjusted walking pace**<br>**I<sup>2</sup> = 61.8%** |  |  | **Walking pace adjusted for BMI**<br>**I<sup>2</sup> = 41.8%** |  |  |
|  | **Beta\* (95% CI)** | **P-value** | **Egger - p** | **Beta\* (95% CI)** | **P-value** | **Egger - p** |
| **MR-IVW** | 0.007 (-0.009, 0.023) | 0.383 |  | 0.006 (-0.006, 0.018) | 0.316 |  |
| **MR-WM** | 0.003 (-0.013, 0.020) | 0.687 | 0.965 | 0.001 (-0.014, 0.017) | 0.852 | 0.978 |
| **MR-RAPS** | 0.007 (-0.003, 0.017) | 0.151 |  | 0.006 (-0.003, 0.016) | 0.185 |  |
| **Part 2: Walking pace on LTL (70 SNPs for walking pace)** |  |  |  |  |  |  |
|  | **Unadjusted walking pace**<br>**I<sup>2</sup> = 72.9%** |  |  | **Walking pace adjusted for BMI**<br>**I<sup>2</sup> = 74.1%** |  |  |
|  | **Beta<sup>†</sup> (95% CI)** | **P-value** | **Egger - p** | **Beta<sup>†</sup> (95% CI)** | **P-value** | **Egger - p** |
| **MR-IVW** | 0.192 (0.077, 0.306) | 0.001 |  | 0.226 (0.063, 0.388) | 0.006 |  |
| **MR-WM** | 0.112 (0.016, 0.209) | 0.023 | 0.563 | 0.110 (-0.021, 0.242) | 0.101 | 0.157 |
| **MR-RAPS** | 0.211 (0.151, 0.271) | 4.44E-12 |  | 0.270 (0.187, 0.353) | 1.51E-10 |  |

Where MR-IVW is the inverse-variance weighted MR which was used as the primary MR method, with MR-WM as the weighted-median MR and MR-RAPS as the robust adjusted profile score MR which were both included as sensitivity analyses. Beta\* is the estimated unit difference in walking pace per 1SD increase in LTL, and Beta<sup>†</sup> is the SD change in LTL per 1 unit difference in walking pace, where a 1 unit increase in self-rated walking pace represents a change in category from slow to steady/average, or from steady/average to brisk pace.

## DISCUSSION

In a large sample of middle-aged adults, we provide evidence that faster self-reported walking pace is associated with longer LTL. In support of the importance of walking pace, using accelerometer-assessed PA we showed that more time habitually spent in higher intensity activities (e.g. brisk walking) had a stronger association with LTL than total activity. Importantly, in bi-directional MR analyses, we also show for the first time that walking pace may be causally associated with LTL, rather than the other way around. Overall, these findings support more intensive habitual movement, such as faster walking pace, as potentially important determinants of LTL and overall health status in humans.

The MR results provide new insights and are important in the context of the current literature, which is limited by a lack of high-quality data and mixed findings from interventional research. The few studies that have been undertaken in this area are generally supportive of our findings. For example, in a non-randomised study, long-term endurance training was associated with reduced LTL erosion compared to healthy non-exercisers [40]. A randomised controlled trial in 68 caregivers (as a model of high exposure to stress) found that 40 minutes of moderate-intensity aerobic exercise 3–5 times per week reduced LTL attrition compared to those in the control group [41]. Further, a recent meta-analysis of case control and intervention studies suggested that across 21 included studies the effect of exercise training produced a moderate effect size (0.7), with exercise associated with longer telomeres [28]; however, associations were no longer statistically significant after accounting for publication bias and stratifying/subgrouping. These findings are also supported by mechanistic studies in animal models showing that chronic exercising increases telomerase and shelterin expression [42-44]. This acts to protect telomere degradation, with an associated reduction of apoptosis and cell-cycle arrest in the myocardium whilst also attenuating age-related erosion of telomeres in hepatocytes and cardiomyocytes [40, 42-44]. Other hypothesised mechanisms linking habitual PA with telomere protection include potential changes in telomerase activity, oxidative stress, inflammation, and decreased skeletal muscle satellite cell content [42, 43, 45].

Interestingly, the suggested causal effect of walking pace on LTL via MR was greater than that suggested by the adjusted observational associations reported in this study, or for exercise intervention studies in middle aged adults [28]. We have previously shown that each additional year in chronological age is associated with a Z-standardised LTL value of -0.024 [34]. Therefore, the difference in LTL between slow and fast walkers suggested by the MR analysis is equivalent to 16 years of age-related difference in LTL, whereas the adjusted observational analysis produced estimates that were equivalent to 2 years of age-related difference. There are several possible explanations for difference. Previously investigated associations between other risk factors and health outcomes, such as blood pressure and lipids, have also reported greater effect sizes from MR than those proposed by observational or interventional research [46-48]. It has been proposed that this difference could be explained by MR measuring lifetime exposure, whereas observational studies measure the exposure at a single point in time, and/or intervention research assesses change over a relative short period of time [49]. However, it has also been cautioned that the association of genetic variants with risk factors may vary by age which could act to inflate MR estimates [50]. Furthermore, differences in the strength of the genetic association with the exposure and outcome may also influence MR estimates [50]. Therefore, whilst MR can help determine causality, the size of the effect should be interpreted with caution and is likely to be greater than the magnitude of change that can be anticipated from any future intervention.

Key strengths of this analysis are the large, contemporary, well-phenotyped cohort with high quality LTL data, and the use of bidirectional MR to examine potential and relative causal effects. However, there are some important limitations. Whilst the self-reported measure of walking pace in UK Biobank has been associated with objectively measured cardiorespiratory fitness [8], it is possible that responses are also influenced by wider factors and personality traits that will not be affected by an intervention to target walking pace *per se*. Nevertheless, analysis of accelerometer data in UK Biobank supported the findings for walking pace, as a measure of habitual movement intensity was associated with longer LTL, whereas there was little evidence of an association for overall movement volume. Although our observational findings were supported by MR, this should be viewed as suggestive of causality, rather than confirmative. The effect sizes and strength of association were attenuated when using the MR-median weighted method, perhaps indicating evidence of potential bias in the walking pace instrument. However, this may be due to reduced power in the method, as our other sensitivity analysis using MR-RAPS was highly significant. Finally, although large in scale, the UK Biobank cohort is healthier than the general population, and the accelerometer sub-study may be subject to some further selection biases (i.e., being measured a median of 5.7 years after UKBB baseline assessment). However, key covariates have been shown to be mostly stable over this time period [51], and risk factor associations have previously been shown to be generalizable to the general population [52].

## Conclusions

A faster habitual walking pace may be causally associated with longer LTL and could help explain some of the beneficial effects of brisk walking on health status. Further research should confirm whether behavioural interventions focused on increasing walking pace or PA intensity act to slow the erosion of LTL. Future work should also elucidate whether these findings simply add support to the use of self-reported walking pace as a measure of overall health status, with a slow walking pace identifying those with potentially accelerated biological ageing, and thus a priority group for other lifestyle/pharmaceutical interventions. Characterising the nature of associations between walking pace and LTL in different population subgroups, particularly those at increased risk of chronic disease or unhealthy ageing, will also be important.

## Data Availability

The UK Biobank data that support the findings of this study can be accessed by researchers on application (https://www.ukbiobank.ac.uk/register-apply/). Variables derived specifically for this study will be returned along with the code to the UK Biobank for future applicants to request.

## DECLARATIONS

### Ethics approval and consent to participate

The UK Biobank study received ethical approval from the North West England Research Ethics Committee (reference 16/NW/0274). Participants gave informed consent before participation.

### Consent for publication

Not applicable

### Availability of data and materials

The UK Biobank resource can be accessed by researchers on application. Variables derived for this study will be returned to the UK Biobank for future applicants to request. No additional data are available.

### Authors’ contributions

PD, AR, VC, CPN, TY, and NJS developed the research question. VC supervised the LTL analysis. PD undertook the observational analyses. CPN, PD, and CM undertook the MR analyses. PD and TY drafted the manuscript. All authors contributed to the interpretation and revised the manuscript for important intellectual content.

## Acknowledgements

We are grateful to the participants of the UK Biobank Study and those who collected and manage the data.

