## Supplemental material for "Causal Associations of Self-Reported Walking Pace with Telomere Length in 405,981 middle-aged adults: a UK Biobank study"

### Accelerometer Methods

A sub-sample of ~100,000 adults were asked to wear the Axivity AX3 wrist-worn accelerometer (Axivity, Newcastle, UK) 24 hours a day for seven days between June 2013 and December 2015 [1]. For each participant, we extracted the accelerometer data (5-second epoch time series) from UK Biobank [1] and converted it to R-format for processing and analysis with GGIR (version 1.11-0, http://cran.r-project.org) [2]. Participants were excluded if they failed calibration (including those not calibrated on their own data), had fewer than three days of valid wear (defined as >16 h per day), or wear data were not present for each 15 min period of the 24 h cycle. Data from n=86,002 UK Biobank participants with valid accelerometer and LTL data, and complete covariate data were included (see **Supplemental Figure S1**).

Accelerometer outcomes, selected to describe total physical activity and its intensity, were:

- ***average acceleration*** over the 24 h day (proxy for total physical activity, m*g*)
- ***intensity gradient*** over 24 h (intensity distribution of physical activity over the day; a high value indicates more time is habitually spent in higher intensity activities, such as brisk walking) [3] – as per the illustration below.

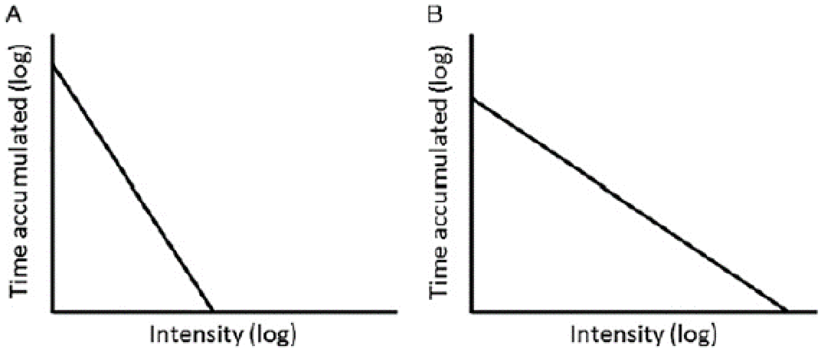


- A. A steeper, more negative (lower) gradient with a higher constant (y-intercept) showing a steep drop in time accumulated with increasing intensity (left)—a 'poorer' intensity profile.
- B. A shallower, less negative (higher) gradient with a lower constant (y-intercept) showing more time spread across the intensity range (right)—a 'better' intensity profile.

**Intensity gradient illustration**. The intensity gradient describes the negative curvilinear relationship between physical activity intensity and the time accumulated at that intensity. It is always negative, reflecting the decrease in time accumulated as intensity increases. Findings for the intensity distribution of PA derived from the accelerometer data were interpreted as being supportive of walking pace, as both relate to habitual movement intensity.

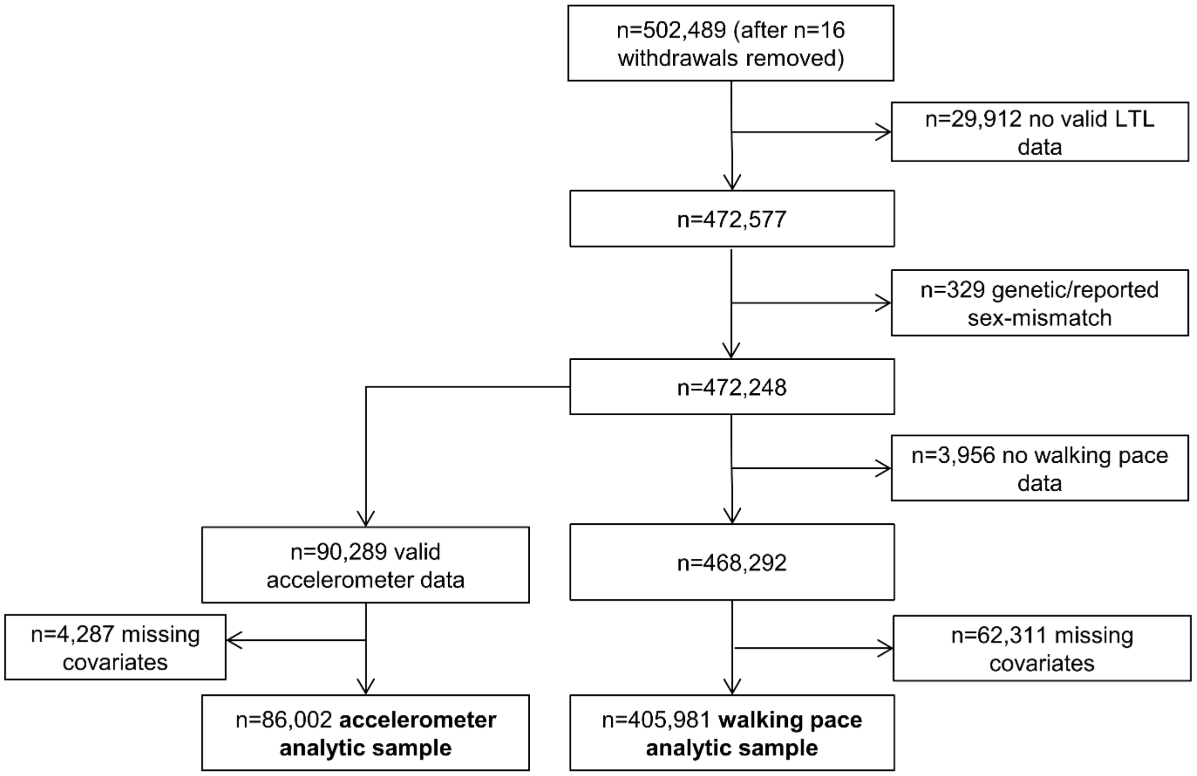


**Supplemental Figure S1:** Flowchart of participant exclusions.

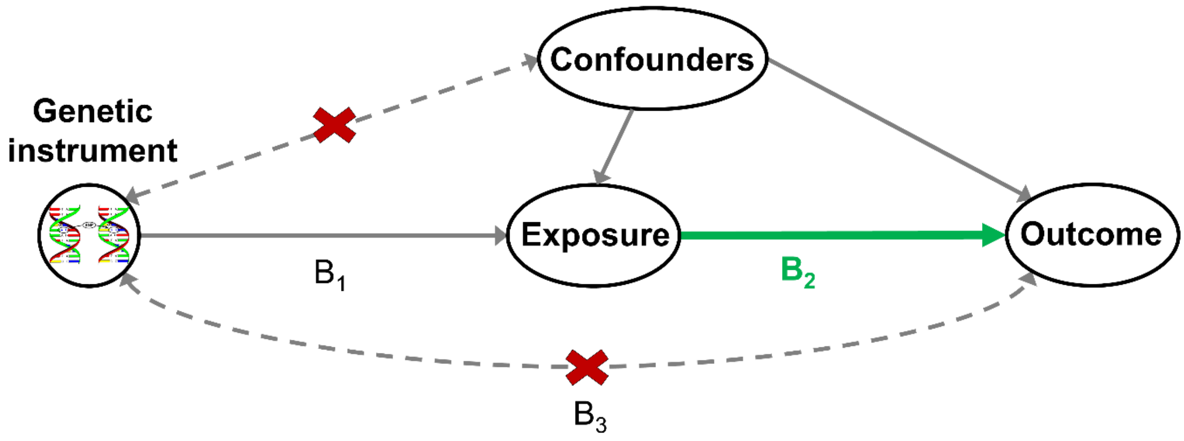


**Supplemental Figure S2:** Simplified causal diagram illustrating Mendelian randomization and its assumptions.

Solid pathway lines are theorized to exist; dashed pathway lines are theorized to be nonsignificant according to model assumptions. $B_2$ indicates the estimated causal relationship ($B_2 = B_1/B_3$). $B_1$ and $B_3$ indicate the estimated direct effects of a genetic variant on the exposure (e.g. walking pace) and outcome (e.g. LTL).

To be valid instrumental variables for the causal association of walking pace on LTL, or vice versa, genetic variants (SNPs) must be: 1) associated with the exposure of interest (e.g. walking pace), 2) independent of factors that confound the association of the exposure and outcome, and 3) associated with the outcome (e.g. LTL) exclusively through their effects on the exposure. If these assumptions are satisfied, the selected SNPs are valid instrumental variables, and their association with disease can be interpreted as a causal effect of the exposure.

**Supplemental Table S1.** Descriptive characteristics at baseline of the accelerometer analytic sample, and by self-reported walking pace.

| Characteristics | Total sample | Slow | Average/steady | Brisk |
|---|---|---|---|---|
| | N=86,002 * | N=3,934 | N=41,305 | N=40,595 |
| Age (years), mean (SD) | 56.16 (7.81) | 58.28 (7.29) | 56.85 (7.73) | 55.24 (7.84) |
| Female gender, n (%) | 48,212 (56.1%) | 2,343 (59.6%) | 23,394 (56.6%) | 22,373 (55.1%) |
| White ethnicity, n (%) | 83,491 (97.1%) | 3,737 (95.0%) | 39,892 (96.6%) | 39,708 (97.8%) |
| **Highest educational level achieved, n (%)** | | | | |
| *No qualification* | 7,057 (8.2%) | 678 (17.2%) | 4,071 (9.9%) | 2,277 (5.6%) |
| *Any other qualification* | 41,492 (48.2%) | 2,051 (52.1%) | 21,097 (51.1%) | 18,268 (45.0%) |
| *Degree level or above* | 37,453 (43.5%) | 1,205 (30.6%) | 16,137 (39.1%) | 20,050 (49.4%) |
| Townsend indicator of multiple deprivation, median (IQR) | -2.46 (-3.83--0.23) | -1.74 (-3.40-1.26) | -2.46 (-3.82--0.23) | -2.53 (-3.86--0.35) |
| In employment, n (%) | 53,183 (61.8%) | 1,584 (40.3%) | 24,308 (58.9%) | 27,244 (67.1%) |
| **Cigarette smoking, n (%)** | | | | |
| *Never* | 49,115 (57.1%) | 1,831 (46.5%) | 22,869 (55.4%) | 24,330 (59.9%) |
| *Previous* | 30,967 (36.0%) | 1,672 (42.5%) | 15,448 (37.4%) | 13,788 (34.0%) |
| *Current* | 5,920 (6.9%) | 431 (11.0%) | 2,988 (7.2%) | 2,477 (6.1%) |
| **Alcohol consumption, n (%)** | | | | |
| *Never or previous* | 4,792 (5.6%) | 498 (12.7%) | 2,373 (5.7%) | 1,886 (4.6%) |
| *< Twice a week* | 38,907 (45.2%) | 2,113 (53.7%) | 19,563 (47.4%) | 17,139 (42.2%) |
| *At least three times a week* | 42,303 (49.2%) | 1,323 (33.6%) | 19,369 (46.9%) | 21,570 (53.1%) |
| **Added salt intake, n (%)** | | | | |
| *Never/rarely* | 51,602 (60.0%) | 2,069 (52.6%) | 24,127 (58.4%) | 25,320 (62.4%) |
| *Sometimes or more frequent* | 34,400 (40.0%) | 1,865 (47.4%) | 17,178 (41.6%) | 15,275 (37.6%) |
| **Oily fish consumption, n (%)** | | | | |
| *More than once a week* | 48,605 (56.5%) | 2,045 (52.0%) | 22,584 (54.7%) | 23,879 (58.8%) |
| Fruit and vegetable intake score, median (IQR) | 2.00 (1.00-2.00) | 1.00 (1.00-2.00) | 2.00 (1.00-2.00) | 2.00 (1.00-3.00) |
| Weekly frequency of red or processed meat intake, median (IQR) | 0.75 (0.50-1.13) | 0.88 (0.50-1.25) | 0.75 (0.50-1.25) | 0.75 (0.50-1.13) |
| **Mean sleep duration, n (%)** | | | | |
| *<7 hours/day* | 18,703 (21.7%) | 1,129 (28.7%) | 8,961 (21.7%) | 8,561 (21.1%) |
| *7-8 hours/day* | 61,878 (71.9%) | 2,252 (57.2%) | 29,465 (71.3%) | 30,087 (74.1%) |
| *>8 hours/day* | 5,421 (6.3%) | 553 (14.1%) | 2,879 (7.0%) | 1,947 (4.8%) |
| **Body mass index, n (%)** | | | | |
| *Normal weight (<25 kg/m2)* | 34,035 (39.6%) | 631 (16.0%) | 12,920 (31.3%) | 20,441 (50.4%) |
| *Overweight (25-30 kg/m2)* | 35,449 (41.2%) | 1,249 (31.7%) | 18,175 (44.0%) | 15,976 (39.4%) |
| *Obese (≥30 kg/m2)* | 16,518 (19.2%) | 2,054 (52.2%) | 10,210 (24.7%) | 4,178 (10.3%) |
| Current prescription of blood pressure or cholesterol medicine, n (%) | 19,627 (22.8%) | 1,793 (45.6%) | 10,781 (26.1%) | 6,965 (17.2%) |
| Diagnosis of diabetes or insulin prescription, n (%) | 2,952 (3.4%) | 466 (11.8%) | 1,627 (3.9%) | 826 (2.0%) |

| | | | | |
|---|---|---|---|---|
| Previous diagnosis of cardiovascular disease, n (%) | 4,187 (4.9%) | 638 (16.2%) | 2,228 (5.4%) | 1,292 (3.2%) |
| Previous diagnosis of cancer, n (%) | 7,085 (8.2%) | 431 (11.0%) | 3,560 (8.6%) | 3,075 (7.6%) |
| Mobility limitation, n (%) | 30,480 (35.4%) | 2,990 (76.0%) | 15,700 (38.0%) | 11,643 (28.7%) |
| Total physical activity (m*g*), mean (SD) | 28.32 (8.41) | 22.65 (7.35) | 27.09 (7.78) | 30.15 (8.64) |
| Intensity gradient, mean (SD) | -2.55 (0.19) | -2.67 (0.20) | -2.57 (0.19) | -2.50 (0.19) |
| **Self-reported walking pace, n (%) *** | | | | |
| *Slow* | 3,934 (4.6%) | - | - | - |
| *Average/steady* | 41,305 (48.0%) | - | - | - |
| *Brisk* | 40,595 (47.2%) | - | - | - |
| *Missing* | 168 (0.2%) | - | - | - |
| Total white blood cell (Leukocyte) count (10^9 cells/Litre), median (IQR) | 6.49 (5.51-7.61) | 7.20 (6.10-8.50) | 6.60 (5.62-7.77) | 6.30 (5.40-7.40) |
| Telomere length (z-score), mean (SD) | 0.000 (1.000) | -0.110 (0.985) | -0.026 (1.004) | 0.037 (0.995) |

* note: n=168 did not have walking pace data in the total (analytic) accelerometer sample (n=86,002) used in subsequent analyses with LTL.

Townsend score, a composite area-level measure of deprivation based on unemployment, non-car ownership, non-home ownership, and household overcrowding; a higher score indicates higher deprivation.

**Supplemental Figure S3.** Baseline exposure distribution and predicted *β*-coefficient (95% CI), relative to the 10th percentile reference of each exposure, for the association of total physical activity (left) and intensity gradient (right) with LTL (n=86,002).

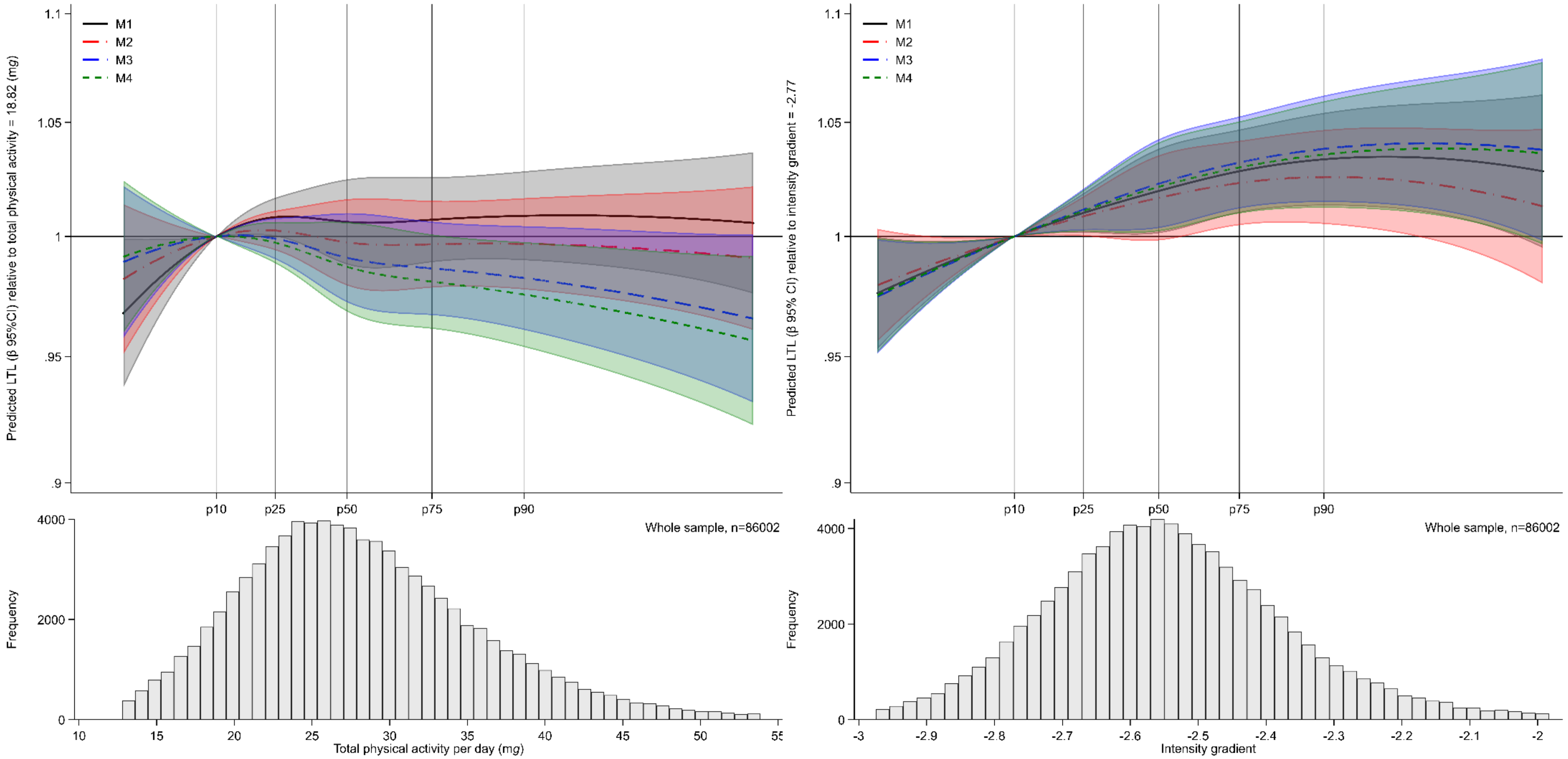


Models were fitted with the use of restricted cubic splines (3 evenly-spaced knots). Predicted LTL *β*-coefficients and histogram data shown for values between the 1st or 99th percentiles of each exposure distribution. The reference values chosen for the exposure are at the 10th percentile (total physical activity=18.82 *mg* ; intensity gradient -2.77), with 25th, 50th, 75th and 90th percentiles also denoted. A higher (less negative) intensity gradient (intensity distribution of physical activity) indicates more time is habitually spent in higher intensity activities (e.g., brisk walking) over a day.

Model 1: is adjusted for age, sex, ethnicity, white blood cell count. Model 2: model 1 additionally adjusted for education level, employment status, Townsend index of deprivation, season of accelerometer wear, fruit and vegetable intake, processed and red meat intake, oily fish intake, regularity of adding salt to food, alcohol intake, smoking status, average sleep duration, blood pressure or cholesterol medication use, diabetes diagnosis or insulin prescription, mobility limitation, and prevalent cardiovascular disease and prevalent cancer. Model 3: model 2 additionally adjusted for either intensity gradient or total physical activity (i.e. mutual adjustment). Model 4: model 3 additionally adjusted for body mass index.
